# Co-circulation and misdiagnosis led to underestimation of the 2015-2017 Zika epidemic in the Americas

**DOI:** 10.1101/19010256

**Authors:** Rachel J. Oidtman, Guido España, T. Alex Perkins

## Abstract

During the 2015-2017 Zika epidemic, dengue and chikungunya – two other viral diseases with the same vector as Zika – were also in circulation. Clinical presentation of these diseases can vary from person to person in terms of symptoms and severity, making it difficult to differentially diagnose them. Under these circumstances, it is possible that numerous cases of Zika could have been misdiagnosed as dengue or chikungunya, or vice versa. Given the importance of surveillance data for informing epidemiological analyses, our aim was to quantify the potential extent of misdiagnosis during this epidemic. Using basic principles of probability and empirical estimates of diagnostic sensitivity and specificity, we generated revised estimates of Zika incidence that accounted for the accuracy of diagnoses made on the basis of clinical presentation with or without molecular confirmation. Applying this method to weekly incidence data from 43 countries throughout Latin America and the Caribbean, we estimated that 1,062,821 (95% CrI: 1,014,428-1,104,794) Zika cases occurred during this epidemic, which is 56.4% (95% CrI: 49.3-62.6%) more than the 679,743 cases diagnosed as Zika. Our results imply that misdiagnosis was more common in countries with proportionally higher incidence of dengue and chikungunya, such as Brazil.

---

Consistent and correct diagnosis is important for the veracity of clinical data used in epidemiological analyses (1–3). Diagnostic accuracy can depend strongly though on the uniqueness of a disease’s symptomatology. On the one hand, diagnosis can be straightforward when there are clearly differentiable symptoms, such as the hallmark rash of varicella (4). On the other hand, with symptoms that are common to many diseases, such as malaise, fever, and fatigue, it can be more difficult to ascertain a disease’s etiology (5–7). Further complicating clinical diagnosis is person-to-person variability in apparent symptoms and their severity (8,9). In many cases, symptoms are self-assessed by the patient and communicated verbally to the clinician, introducing subjectivity and resulting in inconsistencies across different patients and clinicians (10,11).

When they are used, molecular diagnostics are thought to greatly enhance the accuracy of a diagnosis, as they involve less subjectivity and can confirm that a given pathogen is present (12). Even so, molecular diagnostics do have limitations, particularly for epidemiological surveillance. As molecular diagnostics are often not the standard protocol, an infected person first has to present with symptoms in a medical setting and the clinician has to decide to use a molecular diagnostic. This is particularly unlikely to happen for emerging infectious diseases, as clinicians may not be aware of the pathogen or that it is in circulation (13). In this context, molecular diagnostics may also suffer from low sensitivity and specificity, high cost, or unavailability in settings with limited resources (12,14). As a consequence of factors such as these, retrospective analyses of the 2003 SARS outbreak in China identified SARS cases that were clinically misdiagnosed as atypical pneumonia or influenza up to four months before the first laboratory-diagnosed cases of SARS (15).

Challenges associated with disease diagnosis are magnified in scenarios with co-circulating pathogens, particularly when the diseases that those pathogens cause are associated with similar symptoms (16,17). Influenza and other respiratory pathogens, such as *Streptococcus pneumoniae* and respiratory syncytial virus (RSV), co-circulate during winter months in the Northern Hemisphere. The difficulty of correctly ascribing an etiology in this setting is so widely accepted that clinical cases caused by a variety of pathogens are often collated for surveillance purposes as “influenza-like illness” (18). Similar issues occur in malaria-endemic regions (16,19). One study in India found that only 5.7% of commonly diagnosed “malaria-infected” individuals actually had this etiology, while 25% had dengue instead (16).

One set of pathogens with potential for misdiagnosis during co-circulation includes three viruses transmitted by *Aedes aegypti* and *Ae. albopictus* mosquitoes: dengue virus (DENV), chikungunya virus (CHIKV), and Zika virus (ZIKV). Some symptoms of the diseases they cause can facilitate differential diagnosis, such as joint swelling and muscle pain with CHIKV infection (20,21) and a unique rash with ZIKV infection (22,23). Other symptoms, such as malaise and fever, could result from infection with any of these viruses (20–25). In one region of Brazil with co-circulating DENV, CHIKV, and ZIKV, Braga et al. (25) empirically estimated the accuracy of several clinical case definitions of Zika by ground truthing clinical diagnoses against molecular diagnoses. They found that misdiagnosis based on clinical symptoms was common, with sensitivities (true-positive rate) and specificities (true-negative rate) as low as 0.286 and 0.014, respectively.

Although the estimates by Braga et al. (25) provide valuable information about misdiagnosis at the level of an individual patient, they do not address how these individual-level errors might have affected higher-level descriptions of Zika’s epidemiology during its 2015-2017 epidemic in the Americas. The Pan American Health Organization (PAHO) reported 169,444 confirmed and 509,970 suspected cases of Zika across 43 countries between September, 2015 and July, 2017 (26). Meanwhile, PAHO reported 675,476 and 2,339,149 confirmed and suspected cases of dengue and 180,825 and 499,479 confirmed and suspected cases of chikungunya, respectively, during the same timeframe in the same region (27,28). The substantial errors in clinical diagnosis reported by Braga et al. (2017) (25), combined with the large number of cases lacking a molecular diagnosis (26–28), leave open the possibility that a considerable number of cases could have been misdiagnosed during the 2015-2017 Zika epidemic.

Our goal was to quantify the possible extent of misdiagnosis during the 2015-2017 Zika epidemic by leveraging the full extent of passive surveillance data for dengue, chikungunya, and Zika from 43 countries in the Americas in conjunction with empirical estimates of sensitivity and specificity. Our methodology was flexible enough to use either or both of suspected and confirmed cases, given that their availability varied and they both offered information about the incidence of these diseases. To account for variability in diagnostic accuracy, we made use of joint probability distributions of sensitivity and specificity, one for clinical diagnostics and one for molecular diagnostics, informed by empirical estimates. Using this approach, we updated estimates of Zika incidence during its 2015-2017 epidemic across the Americas.

## Methods

To quantify the degree of misdiagnosis during the Zika epidemic, we leveraged the full extent of passive surveillance data on Zika, dengue, and chikungunya for 43 countries in the Americas and formulated a Bayesian model of the passive surveillance observational process. Our observation model was informed by the observed proportion of Zika and empirically estimated misdiagnosis rates (Fig. 1). We used the model to generate revised estimates of the number of Zika cases that occurred during the 2015-2017 Zika epidemic across the Americas (Fig. 1).

**Figure 1.**
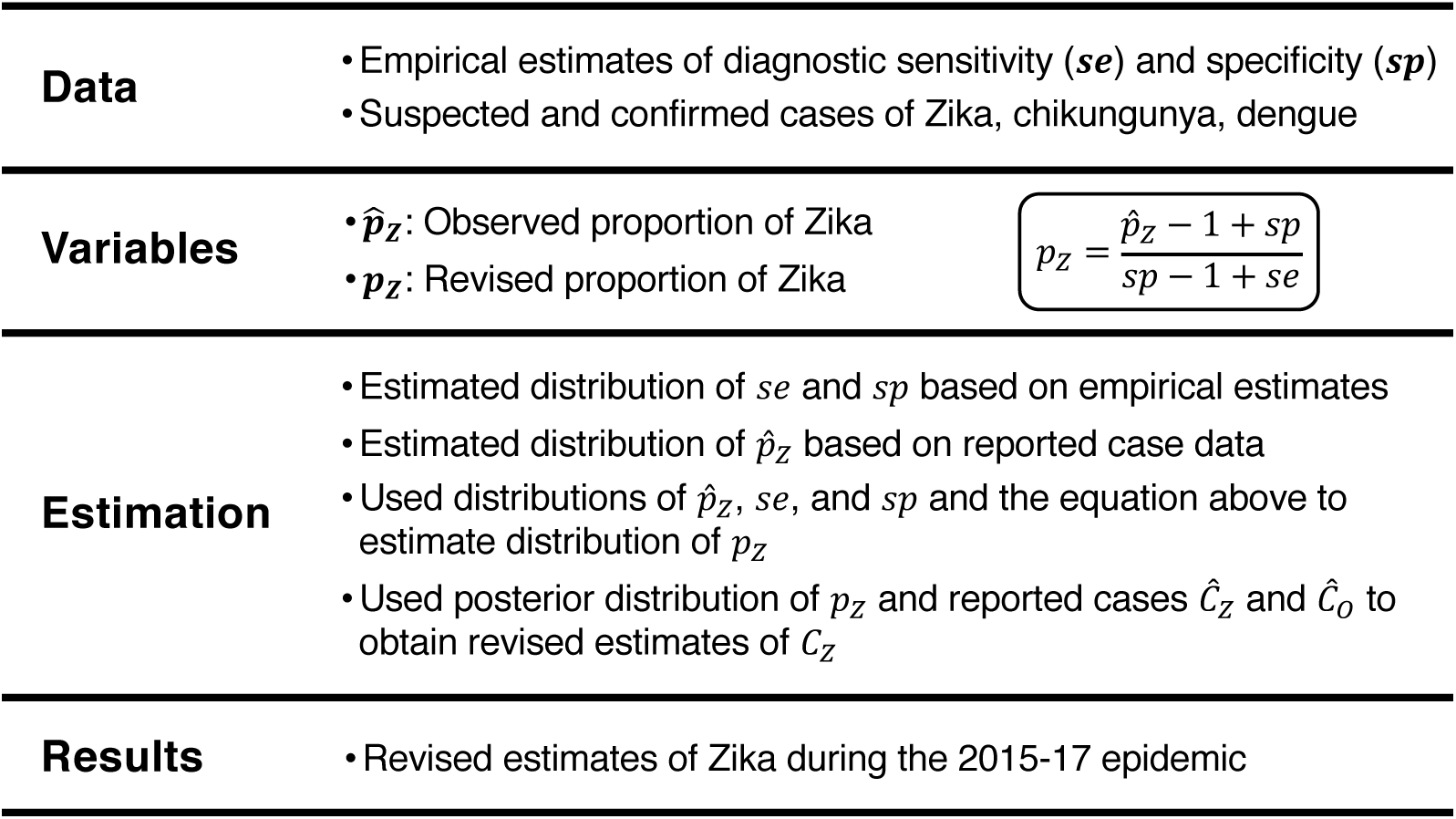
Overview of approach.

### Data

We used suspected and confirmed case data for dengue, chikungunya, and Zika from PAHO for 43 countries in the Americas. We differentiated between confirmed and suspected cases on the basis of laboratory diagnosis versus clinical diagnosis (29). For confirmed and suspected cases of chikungunya, we used manual extraction and text parsing algorithms in perl to automatically extract data from epidemiological week (EW) 42 of 2013 through EW 51 of 2017 (28). For confirmed and suspected cases of Zika, we used the skimage (30) and numpy (31) packages in Python 3.6 to automatically extract incidence data from epidemic curves for each country from PAHO from EW 39 of 2015 to EW 32 of 2017 (26). For confirmed and suspected cases of dengue, we downloaded weekly case data available from PAHO from EW 42 week of 2013 to EW 51 of 2017 (27). We restricted analyses to EW 42 of 2015 (the beginning of the fourth quarter of 2015) to EW 32 of 2017 (the last week with Zika data in our dataset) to focus our analysis on the timeframe of the Zika epidemic (Fig. 2).

**Figure 2.**
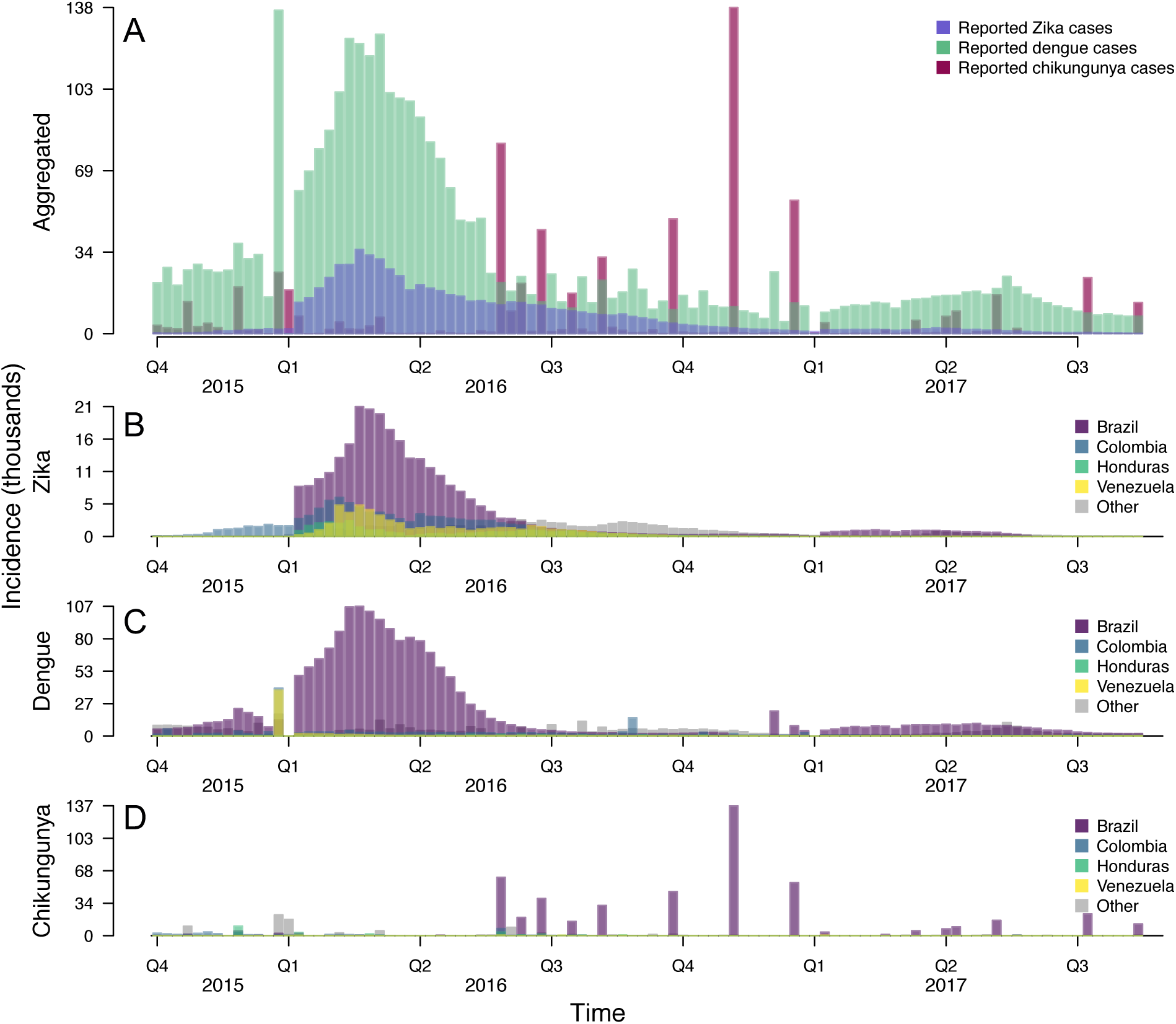
Weekly incidence of Zika, dengue, and chikungunya across the Americas with suspected and confirmed cases combined. A: Incidence aggregated across the entire region. Country-specific reports of B: Zika, C: dengue, and D: chikungunya for Brazil, Colombia, Honduras, Venezuela, and all other countries aggregated as “Other.” These four countries were chosen for visual purposes, as they had the highest total Zika incidence during the epidemic period.

### Probabilistic estimates of sensitivity and specificity

Due to variability in the sensitivity and specificity of different diagnostic criteria, we treated *se* and *sp* as jointly distributed random variables informed by empirical estimates. To describe variability in misdiagnosis for molecular diagnostic criteria, we included two empirical estimates of molecular sensitivity and specificity that were found using ZIKV RT-PCR on a panel of samples with known RNA status for ZIKV, DENV, CHIKV, or yellow fever virus (32). To describe variability in misdiagnosis for clinical diagnostic criteria, we included six empirical estimates of sensitivity and specificity that were measured in a region of Brazil with co-circulating ZIKV, DENV, and CHIKV (25). These empirical estimates of sensitivity and specificity were derived by clinically diagnosing a patient with Zika, dengue, or chikungunya based on different clinical case definitions, and then ground truthing against the case’s etiology determined by RT-PCR (25). We used the sample mean, ***μ***, and sample variance-covariance matrix, **Σ**, for the molecular and clinical misdiagnosis rates as the mean and covariance in two independent, multivariate normal distributions, such that

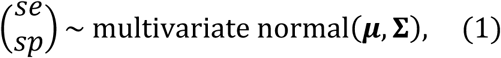

for each of the molecular and clinical diagnostic distributions.

### Probabilistic estimates of the proportion of Zika

We used the proportion of cases that were diagnosed as confirmed or suspected Zika, 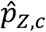 and 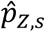, where *c* and *s* refer to confirmed and suspected cases and the hat notation refers to observed data. Rather than using the point estimate for 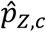 or 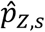, we made use of Bayesian posterior estimates of these variables obtained directly from reported Zika cases, *Ĉ*_*Z,c*_ and *Ĉ*_*Z,s*_, and reported dengue and chikungunya cases, *Ĉ*_*O,c*_ and *Ĉ*_*O,s*_, using the beta-binomial conjugate relationship (33). This assumed that the number of Zika cases was a binomial draw from the total number of cases of these three diseases, with a beta-distributed probability of success, 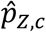 or 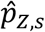. We assumed uninformative priors on 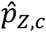 and 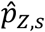; i.e., 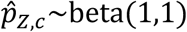 and 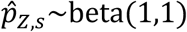. Therefore, 1 + *Ĉ*_*Z,c*_ and 1 + *Ĉ*_*Z,s*_ were the alpha parameters of the two beta distributions and 1 + *Ĉ*_*O,c*_ and 1 + *Ĉ*_*O,s*_ were the beta parameters of the two beta distributions. For confirmed cases, this resulted in a posterior estimate of

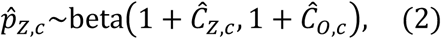

and for suspected cases,

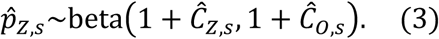

### Observation model of misdiagnosis

To calculate the updated proportion of Zika among the total of Zika, dengue, and chikungunya cases, *p*_*Z*_, we mathematically related 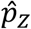 to *p*_*Z*_ using diagnostic sensitivity and specificity, such that

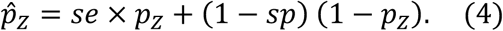

We then rearranged Eq. 4 to solve for

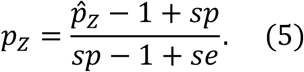

From Eq. 5, we determined two constraints for how *se, sp*, and 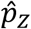 can relate to one another. The first was 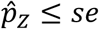, which follows from 0 ≤ *p*_*Z*_ ≤ 1, or 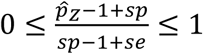, and then simplifying the inequality. The second was *se* + *sp* ≠ 1, as this would lead to zero in the denominator of Eq. 5. Eqs. 4, 5, and subsequent constraints were applied independently to confirmed and suspected cases.

We used samples of *p*_*Z,c*_ and *p*_*Z,s*_ estimated from Eq. 5 to define a single distribution of *p*_*Z*_. As estimates of *p*_*Z,c*_ and *p*_*Z,s*_ were between 0 and 1, we approximated beta distributions for each using the fitdistr function in the MASS package in R (34) fitted to posterior samples of *p*_*Z,c*_ and *p*_*Z,s*_. We then reconciled differences in estimates of *p*_*Z,c*_ and *p*_*Z,s*_ with an estimate of *p*_*Z*_, defined as

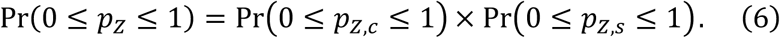

We then applied *p*_*Z*_ to *Ĉ*_*Z,c*_ + *Ĉ*_*Z,s*_ + *Ĉ*_*O,c*_ + *Ĉ*_*O,s*_ to obtain revised Zika cases, *C*_*Z*_, and dengue and chikungunya cases, *C*_*O*_.

### Applying the observation model

To apply our observation model to empirical data, we first drew 1,000 samples from the beta distributions of 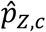 and 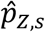 and 1,000 samples from the multivariate normal distributions describing sensitivities and specificities of molecular and clinical diagnostics. We applied our observation model to four distinct scenarios with different spatial and temporal aggregations to assess the sensitivity of our results to different ways of aggregating incidence data: country-specific, temporal data (4,214 data points); country-specific, cumulative data (43 data points); region-wide, temporal data (98 data points); and region-wide; cumulative data (1 data point). Under each of these scenarios, we quantified posterior distributions of *p*_*Z*_, drew 1,000 Monte Carlo samples of *p*_*Z*_, and obtained distributions of *C*_*Z*_ and *C*_*O*_.

## Results

### Illustrative example

We constructed a simple example with two generic diseases, A and B, to illustrate the relationship between reported cases and revised cases under different misdiagnosis scenarios. For these generic diseases, we varied the total cases of A and B such that the proportion of cases diagnosed as A, 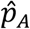, varied from high to low. We used combinations of sensitivity and specificity that spanned all combinations of low, intermediate, and high misdiagnosis scenarios. Using the same methods applied to reallocate Zika, dengue, and chikungunya cases, we revised estimates of incidence of disease A in light of misdiagnosis with disease B.

Incidence of disease A was not revised when sensitivity and specificity were both low (Fig. 3, bottom left), which was due in some cases to the constraint of 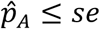 not being met and in other cases to the sum of sensitivity and specificity equaling 1. When 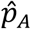 was high (Fig. 3, pink lines), revised incidence of A was similar to observed incidence of A, as only high sensitivities were possible across a range of specificities (Fig. 3, top row). With high sensitivities (Fig. 3, top row), misdiagnosis only occurred with B misdiagnosed as A. When 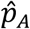 was low (Fig. 3, purple lines), revised incidence of A was higher than observed incidence, as only high specificities were possible across a range of sensitivities (Fig. 3, right column). With high specificities (Fig. 3, left column), misdiagnosis only occurred with A misdiagnosed as B. When 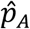 was intermediate (Fig. 3, green lines), misdiagnosis occurred both ways, as a range of sensitivity and specificity values were possible. This resulted in scenarios in which incidence of A was higher or lower than the observed incidence.

**Figure 3.**
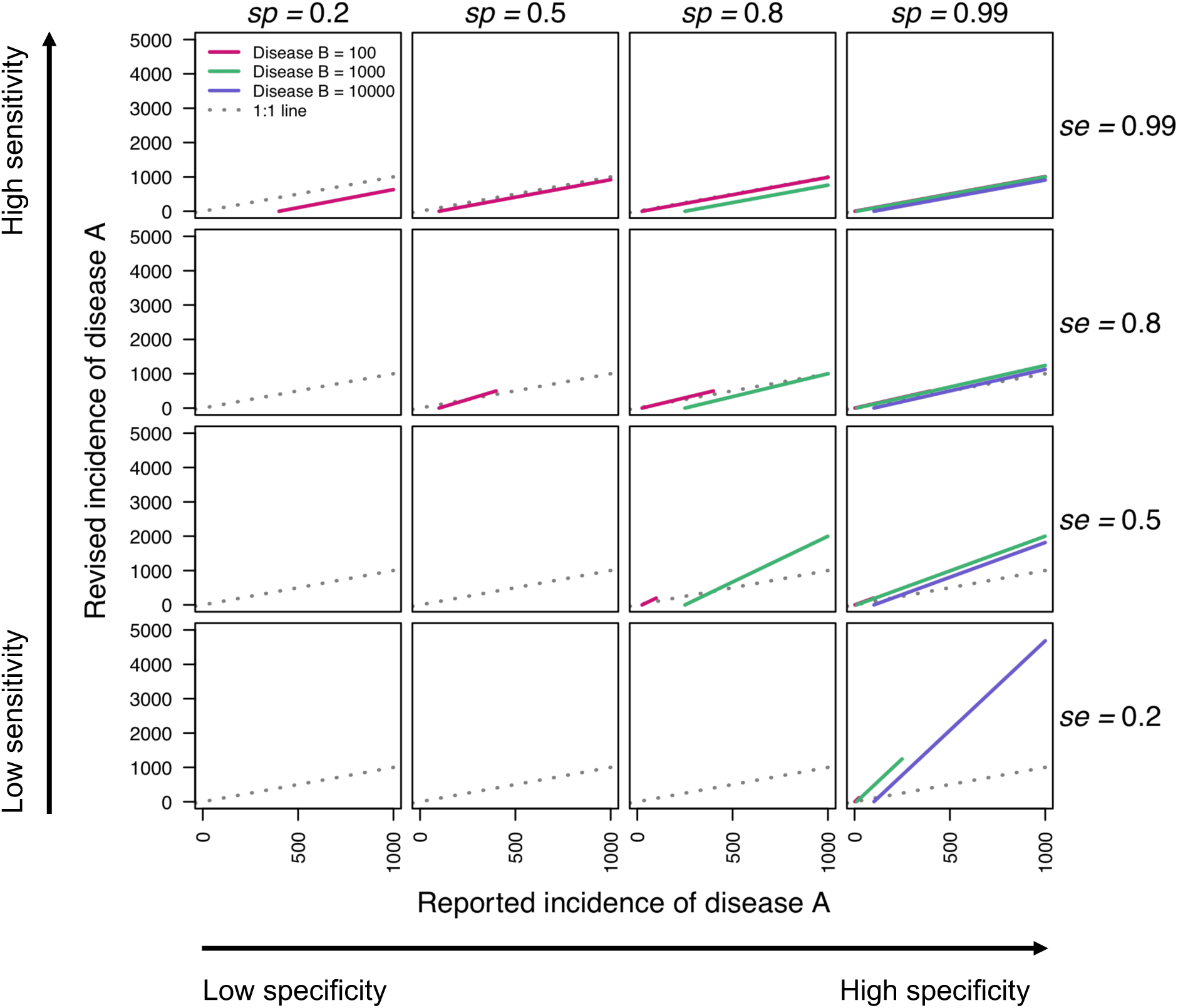
Relationships between reported and revised incidence of disease A under different misdiagnosis scenarios with disease B. Sensitivity and specificity values span 0.2, 0.5, 0.8, and 0.99 across rows and columns. Colors denote different values of observed incidence of disease B. The gray line is the 1:1 line, which separates when revised incidence of disease A is higher than reported (above) and when revised incidence of disease A is lower than reported (below). Plots with no lines indicate that a constraint was broken (se + sp ≠ 1 or 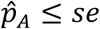). Lines only span portions of the x-axis under which revised incidence of disease A is positive.

### Misdiagnosis through time

We estimated the revised proportion of Zika cases among all cases of Zika, dengue, and chikungunya at each time point for each country. As 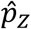 was low early in the epidemic, we estimated that there were 113,459 (95% CrI: 74,595-148,575) disease episodes caused by ZIKV that were misdiagnosed as dengue or chikungunya cases in the fourth quarter of 2015, prior to the start of reporting of Zika in most countries (Fig. 4). Then, as Zika incidence increased and peaked in 2016, the intensity of misdiagnosis increased (Fig. 4), but the direction of misdiagnosis differed by country, depending on how much 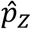 increased. Drastic jumps and dips in the number of Zika cases misdiagnosed as dengue or chikungunya (Fig. 4) was a consequence of chikungunya cases not being reported on a continuous basis (Fig. 2D).

**Figure 4.**
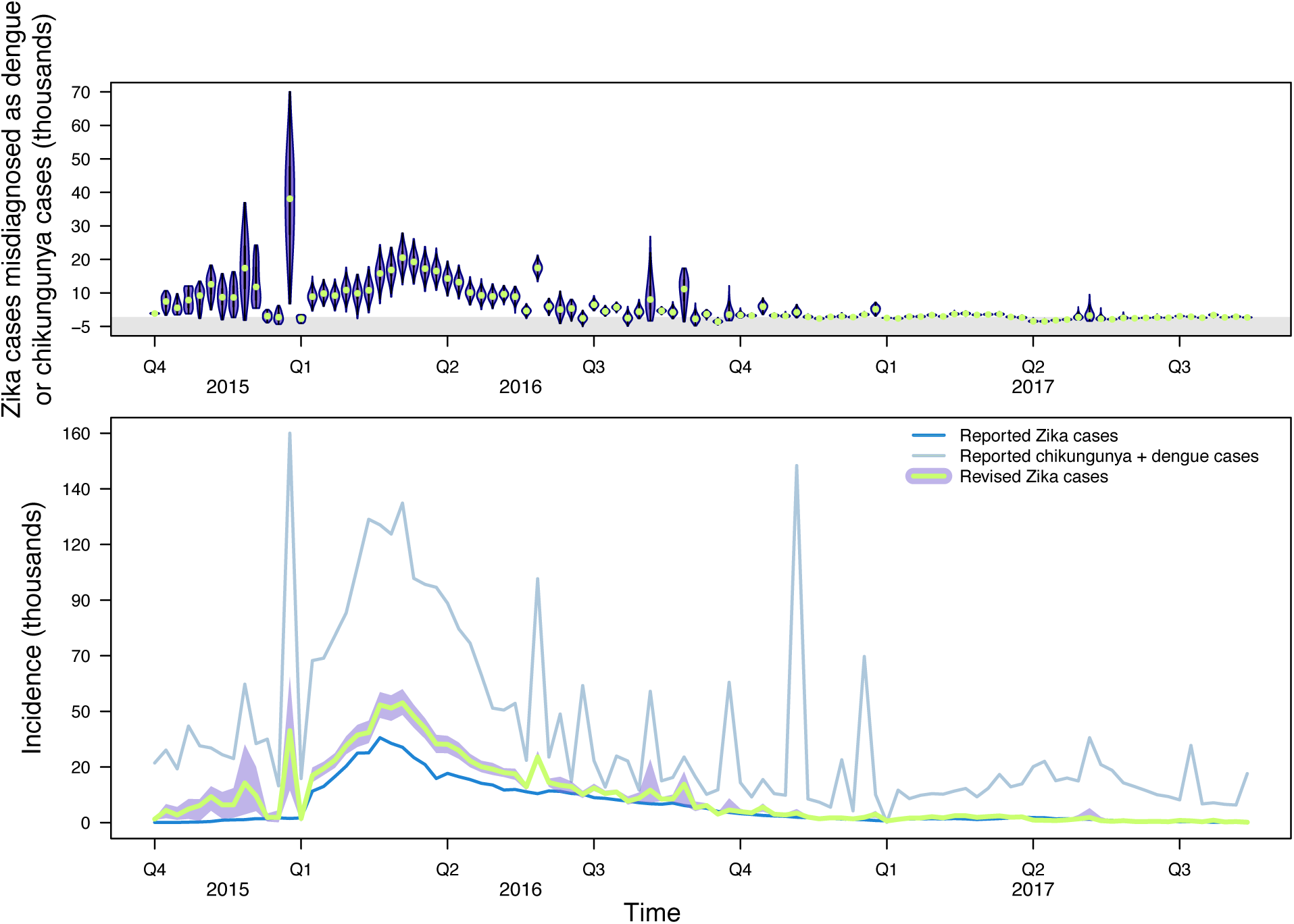
Estimates of revised Zika incidence after accounting for misdiagnosis with dengue and chikungunya. Top: Violin plots of the number of Zika cases that were misdiagnosed as chikungunya or dengue cases on a country-level that were then aggregated across the region for visualization. Estimates above zero indicate that there were more Zika cases than observed and estimates below zero (gray region) indicate there were fewer Zika cases than observed. Bottom: Reported Zika, dengue, and chikungunya incidence alongside revised estimates of Zika incidence and associated uncertainty. Purple band is 95% CrI and green line is median estimate.

### Revising cumulative estimates of the epidemic

We aggregated revised Zika incidence to estimate the cumulative size of the epidemic and to compare our estimate to that based on surveillance reports. In countries and territories with relatively high Zika incidence (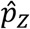 close to 1), such as Suriname, Martinique, and the U.S. Virgin Islands, our revised estimates of *p*_*Z*_ closely matched 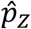 (Fig. 5, bottom). In countries with relatively low Zika incidence (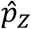 close to 0), such as Brazil and Bolivia, our revised estimates of *p*_*Z*_ were higher than 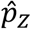 (Fig. 5, bottom). In those countries that reported no Zika incidence (i.e., 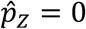), such as Bermuda and Chile, our estimates of *p*_*Z*_ were much more uncertain (Fig. 5, bottom).

**Figure 5.**
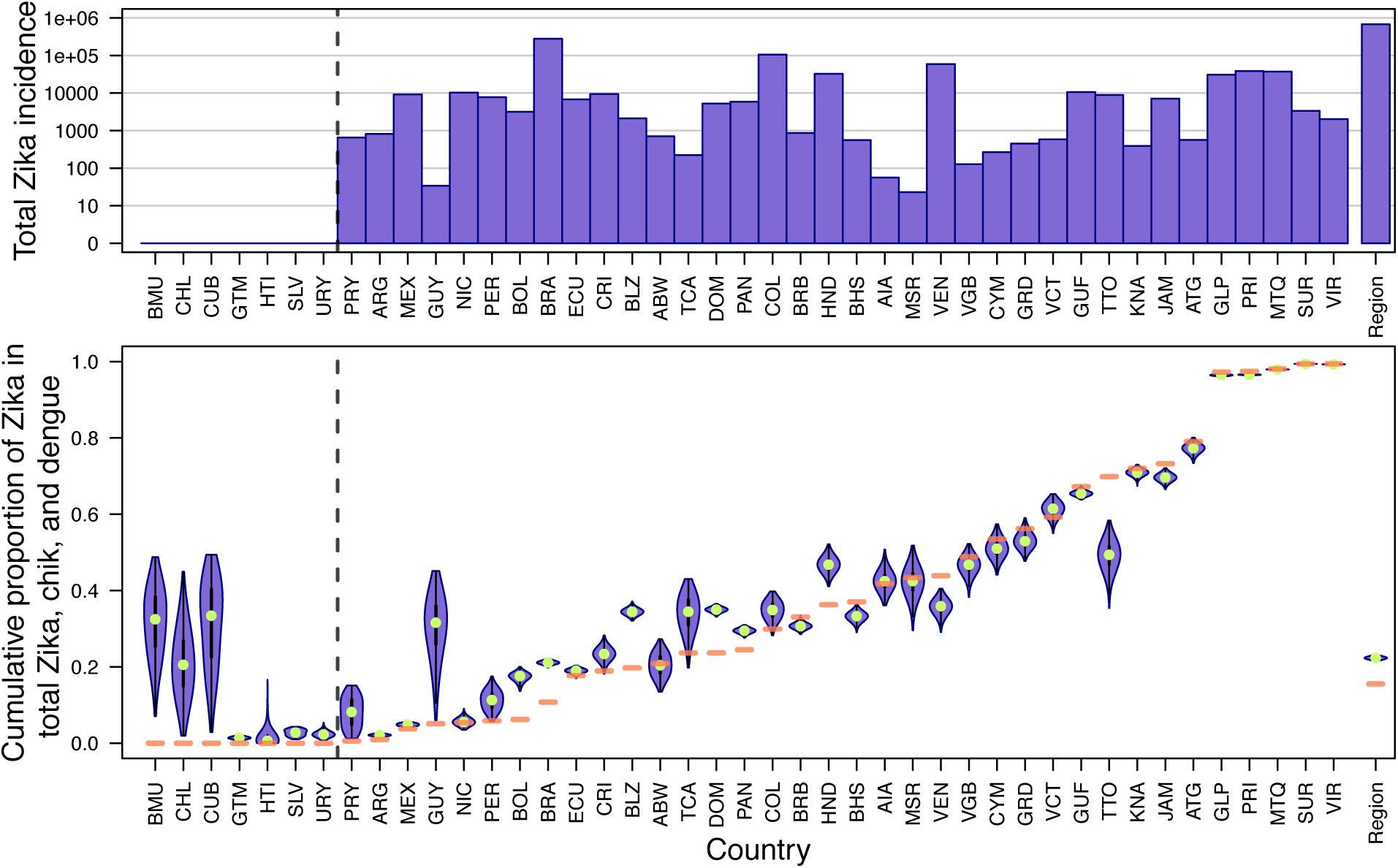
Revised estimates of cumulative p_Z_ versus empirical 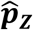 by country. Countries to the left of the dotted line reported zero confirmed or suspected Zika cases according to PAHO. Top: Total Zika incidence by country on a log_10_ scale. Bottom: Violin plots of cumulative p_Z_, with empirical 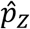 indicated with a horizontal orange line. The distributions of p_Z_ were estimated using temporally disaggregated data for each country and were then were aggregated for visualization. Region-wide estimates are shown farthest to the right.

According to the PAHO reports that we used, the Zika epidemic totaled 679,414 confirmed and suspected cases throughout 43 countries in the Americas. When we accounted for misdiagnosis among Zika, dengue, and chikungunya, we estimated that the Zika epidemic totaled 1,062,821 (95% CrI: 1,014,428-1,104,794) cases across the Americas.

### Estimates of epidemic size using different aggregations of data

We applied our observation model to different temporal and spatial aggregations of the PAHO data, wherein we used temporal incidence data for the region as a whole (Fig. S4), cumulative incidence data for each country (Fig. S5), or cumulative incidence data for the region as a whole (Table 2). When using temporal incidence data for the region as a whole, our estimate of the overall size of the Zika epidemic was 1,073,593 (95% CrI: 1,049,660-1,101,840). Under this spatially aggregated scenario, the majority of misdiagnosis occurred during the height of the epidemic (Fig. S2). In our analysis using cumulative incidence data for each country, our estimate of the overall size of the epidemic was 844,623 (95% CrI: 724,421-957,294), with country-specific estimates of *p*_*Z*_ not well-aligned with 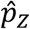 (Fig. S3). When using cumulative incidence for the region as a whole, our estimate of the overall size of the Zika epidemic was 227,568 (95% CrI: 139,782-319,684).

**Table 1.** Empirical sensitivity and specificity values used to inform distributions of sensitivity and specificity for confirmed and suspected cases.

| Diagnostic criteria | Sensitivity | Specificity | Reference |
| --- | --- | --- | --- |
| Molecular | 0.87 | 0.95 | (32) |
| Molecular | 0.82 | 0.96 | (32) |
| Clinical | 0.813 | 0.109 | (25) |
| Clinical | 1.0 | 0.014 | (25) |
| Clinical | 0.583 | 0.519 | (25) |
| Clinical | 0.809 | 0.580 | (25) |
| Clinical | 0.756 | 0.635 | (25) |
| Clinical | 0.286 | 0.973 | (25) |

**Table 2.**
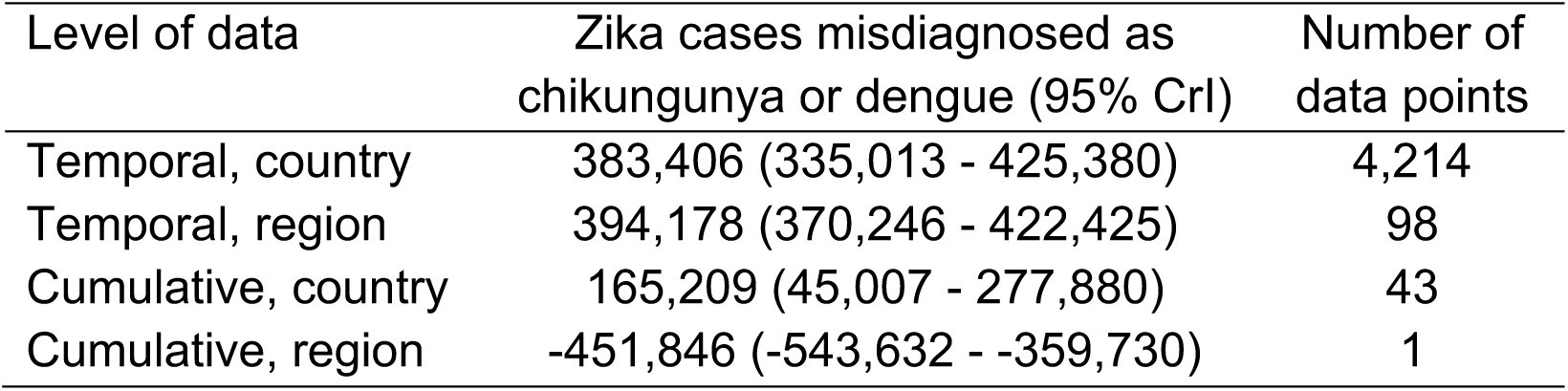
Revised estimates of cumulative Zika incidence across the Americas using different spatial and temporal aggregations of incidence data.

| Level of data | Zika cases misdiagnosed as<br>chikungunya or dengue (95% CrI) | Number of<br>data points |
| --- | --- | --- |
| Temporal, country | 383,406 (335,013 - 425,380) | 4,214 |
| Temporal, region | 394,178 (370,246 - 422,425) | 98 |
| Cumulative, country | 165,209 (45,007 - 277,880) | 43 |
| Cumulative, region | -451,846 (-543,632 - -359,730) | 1 |

## Discussion

We leveraged empirical estimates of sensitivity and specificity for both clinical and molecular diagnostics to revise estimates of the 2015-2017 Zika epidemic in 43 countries across the Americas. After applying our methods to data from PAHO, we found that more than 300,000 disease episodes diagnosed as chikungunya or dengue from September, 2015 through July, 2017 may have been caused by ZIKV instead. Our revised estimates of the Zika epidemic suggest that the epidemic was more than 50% larger than case report data alone would suggest. Additionally, our estimates show that nearly a third of these instances of misdiagnosis occurred in 2015, prior to many countries reporting Zika cases to PAHO (26). An illustrative example of our method showed that these results were driven by the relative incidence of Zika and the two other diseases. Hence, differences in our results over time, across countries, and with respect to level of data aggregation resulted from differences in relative incidence of Zika and these other diseases across the different ways of viewing the data that we considered.

Some countries appeared to have higher Zika incidence than surveillance data alone suggest, such as Brazil and Bolivia, while others appeared to have lower Zika incidence than surveillance data alone suggest, such as Venezuela and Jamaica. In Brazil and Bolivia, our country-specific cumulative estimates of the epidemic were 90% and 180% larger than case report totals, respectively. In Venezuela and Jamaica, our country-specific estimates of the epidemic were 29% and 17% smaller than case report totals, respectively. These differences across countries can be explained by differences in the proportions of suspected Zika cases, 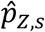, through time. In Brazil and Bolivia, 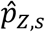 was less than 0.2 at nearly every time point, whereas it mostly ranged 0.2-0.8 in Venezuela and Jamaica. When 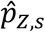 was low, as in Brazil and Bolivia, the constraint that 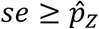 allowed sensitivities to span a larger range, including lower sensitivities that would have resulted in the inference that more cases diagnosed as dengue or chikungunya were caused by ZIKV. When 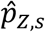 was moderate to high, as in Venezuela and Jamaica, the constraint that 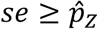 limited sensitivities to higher values, resulting in the inference that fewer cases diagnosed as dengue or chikungunya were caused by ZIKV. Similarly, because of a trade-off between sensitivity and specificity for clinical diagnoses, these constraints on sensitivity also imposed constraints on specificity.

Using different aggregations of data in our analysis led to different conclusions in multiple respects. Using cumulative data for the region as a whole led to the inference that the Zika epidemic was smaller than suggested by surveillance data, whereas using cumulative data at a country level led to the inference that the epidemic was larger than suggested by surveillance data, but with variation across countries. Using temporally explicit data led to the inference that the epidemic was even larger, regardless of whether data was aggregated at a country or regional level. Overall, these similarities and differences suggest greater consistency temporally than spatially in the relative incidence of Zika, chikungunya, and dengue across countries. At least in the case of an emerging disease such as Zika, this suggests that it may be most important to prioritize temporal data when inferring patterns of misdiagnosis. With respect to the timing of inferred misdiagnoses, there were more visible differences between scenarios in which temporal data were aggregated at a country or regional level. When temporal data were aggregated at a country level, we inferred that the majority of misdiagnosis occurred prior to 2016. When temporal data were aggregated at the regional level, we inferred that the majority of misdiagnosis occurred during the epidemic in 2016.

Our observation model incorporated basic features of how passive surveillance data for diseases caused by multiple, co-circulating pathogens are generated, including the potential for misdiagnosis and differences in misdiagnosis rates by data type. With respect to other features of how data such as these are generated, there were some limitations of our approach. First, we did not specify a process model of transmission dynamics. For example, a multiple-pathogen transmission model could be fitted to passive surveillance data on Zika, dengue, and chikungunya, using our observation model to relate the model’s predictions to the data (33). Accounting for misdiagnosis in this way could improve a transmission model’s ability to make inferences about drivers of transmission (35–37) or interactions among pathogens (38,39). Second, we aggregated chikungunya and dengue case data, meaning that we were unable to explore the potential for differences in the extent to which misdiagnosis occurs between each of these diseases and Zika. If additional studies resolve differences in diagnostic sensitivity and specificity of Zika compared to each of these diseases separately, our observation model could be extended to account for this. Third, our observation model relied on a limited set of empirical estimates of diagnostic sensitivity and specificity. Given that the use of different diagnostics could vary spatially or temporally, as could their sensitivities and specificities (40–42), incorporating more detailed information about diagnostic use and characteristics could improve future estimates using our observation model.

Although passive surveillance data has been central for understanding many aspects of the 2015-2017 Zika epidemic, our finding that there may have been 56% more Zika cases than described in PAHO case reports underscores the need to consider the observation process through which passive surveillance data is collated. Here, we accounted for misdiagnosis in the observation process to revise estimates of the passive surveillance data on which numerous analyses depend (35,43,44). The advancements made here contribute to our understanding of which pathogen may be circulating at a given time and place. By better accounting for the etiology of reported cases, it could become more feasible to implement pathogen-specific response measures, such as proactively testing pregnant women for ZIKV during a Zika epidemic (45,46). Given the potential for synchronized epidemics of these and other co-circulating pathogens in the future (47), continuing to develop methods that disentangle which pathogen is circulating at a given time will be important in future epidemiological analyses based on passive surveillance data.

## Data Availability

Data available on GitHub at https://github.com/roidtman/zika_misdiagnosis.

## Acknowledgments

RJO acknowledges support from an Arthur J. Schmitt Leadership Fellowship in Science and Engineering and an Eck Institute for Global Health Fellowship. GE acknowledges support from Grant TL1TR00107 (A Shekhar, PI) from the National Institutes of Health, National Center for Advancing Translational Sciences, Clinical and Translational Sciences Award (https://www.indianactsi.org). TAP acknowledges support from a Young Faculty Award from the Defense Advanced Research Projects Agency (D16AP00114), a RAPID Award from the National Science Foundation (DEB 1641130), and Grant P01AI098670 (T Scott, PI) from the National Institutes of Health, National Institute of Allergy and Infectious Disease. The authors thank Dr. Ann Raiho for helpful comments on the manuscript.

## SUPPLEMENTAL FIGURES

**Figure S1.**
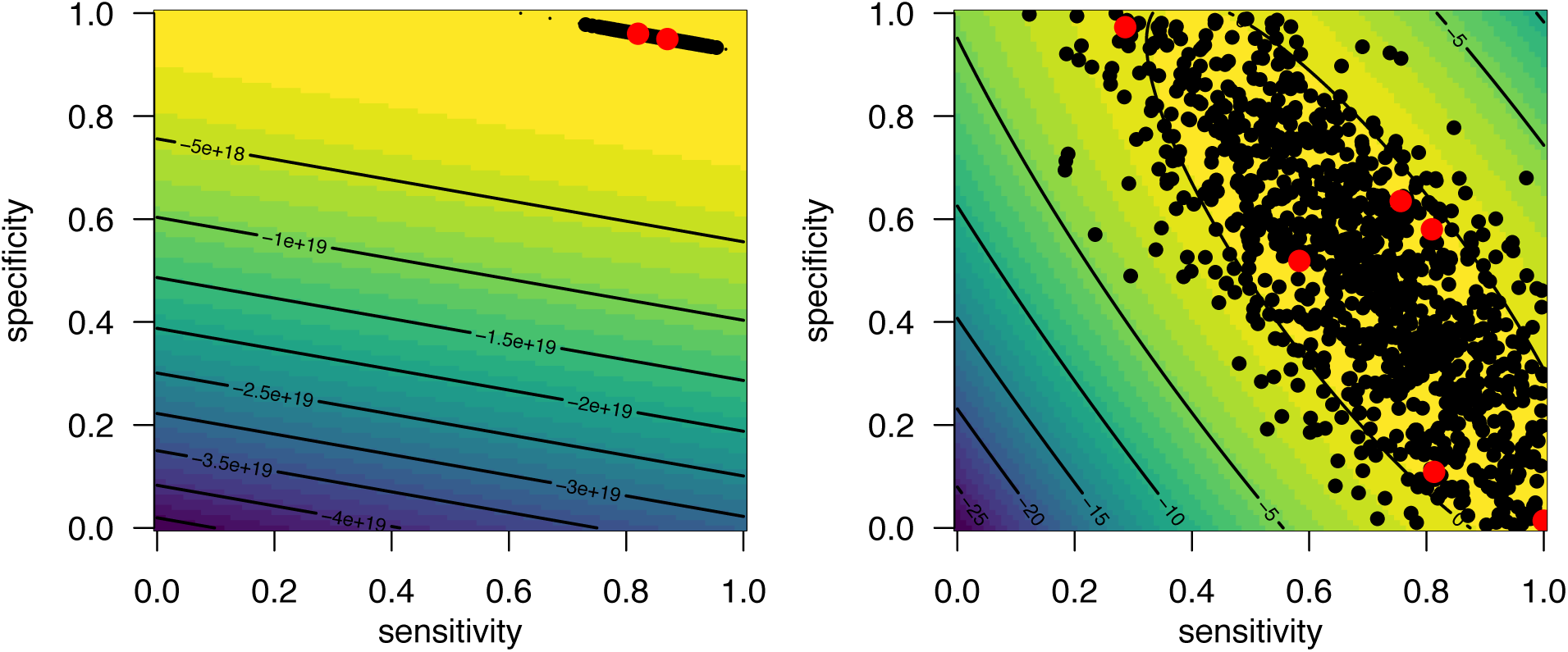
Multivariate normal distributions fitted to empirical sensitivities and specificities of molecular diagnostics (left) and empirical sensitivities and specificities of clinical diagnostics (right). Red points are empirical estimates, and black points are samples from the multivariate normal distributions. On the probability surface, yellow indicates high probability and navy indicates low probability.

**Figure S2.**
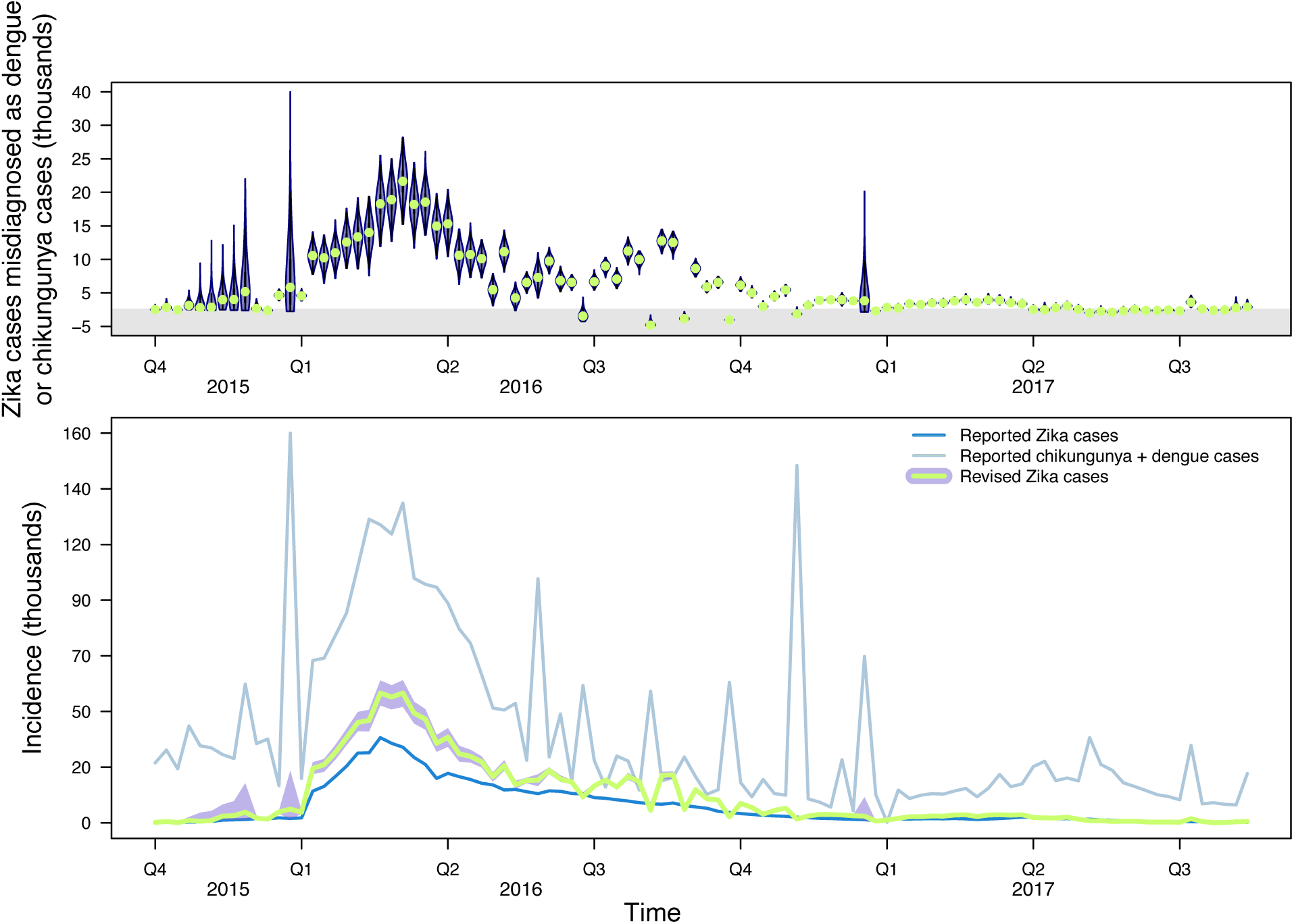
Estimates of Zika incidence after accounting for misdiagnosis using spatially aggregated data. Top: Violin plots of the number of Zika cases that were misdiagnosed as chikungunya or dengue cases. Estimates above zero indicate there were more Zika cases than perceived and estimates below zero (gray region) indicate there were fewer Zika cases than perceived. Bottom: Reported Zika and dengue and chikungunya incidence alongside revised estimates of Zika incidence with associated uncertainty. Purple band is 95% CrI and green line is median estimate.

**Figure S3.**
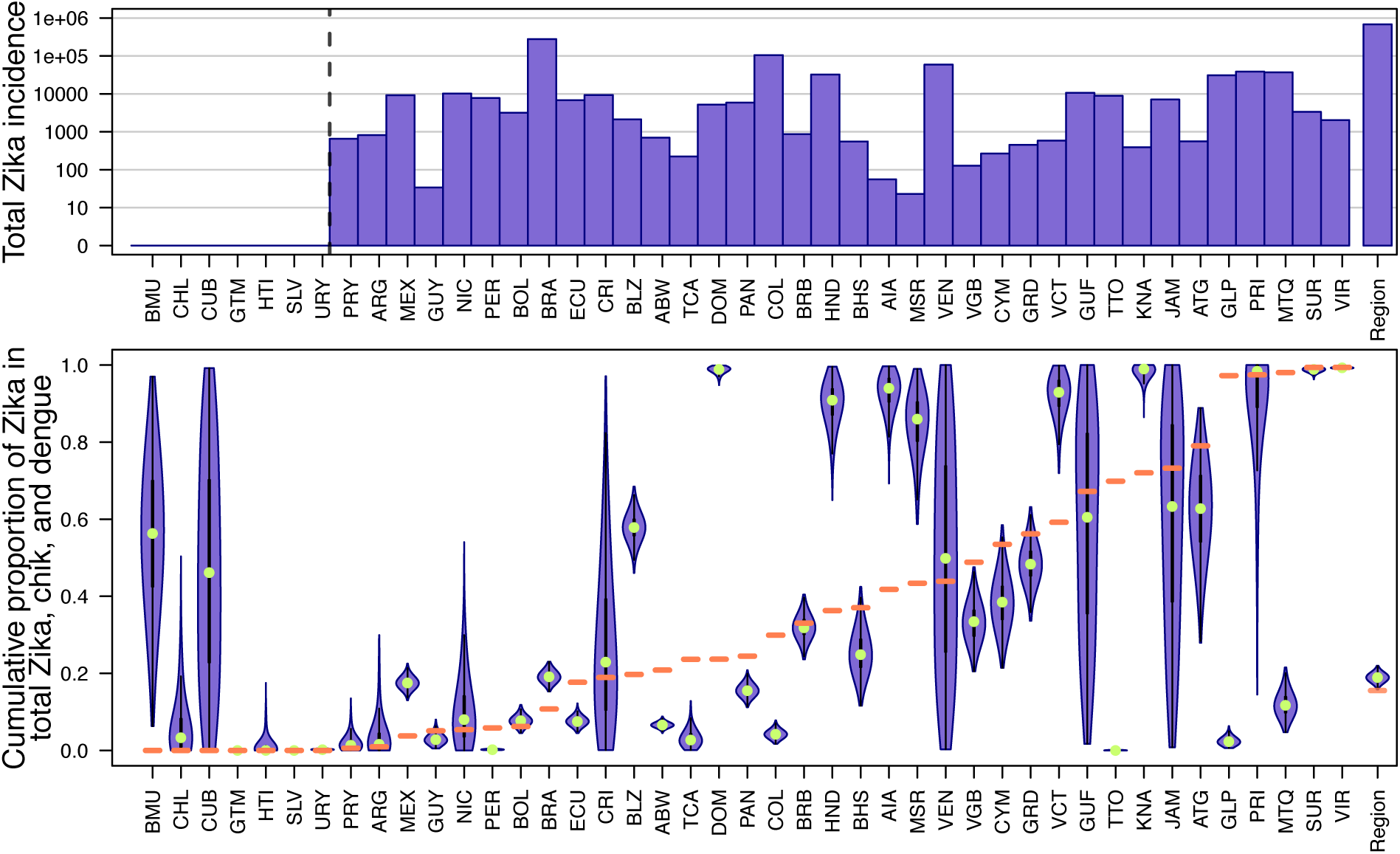
Revised estimates of cumulative p_Z_ versus empirical 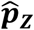 by country using cumulative data only. Countries to the left of the dotted line reported zero confirmed or suspected Zika cases according to PAHO. Top: Total Zika incidence by country on a log_10_ scale. Bottom: Violin plots of cumulative p_Z_ with empirical 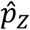 indicated with a horizontal orange line. Region-wide estimate are shown farthest to the right.

**Figure S4.**
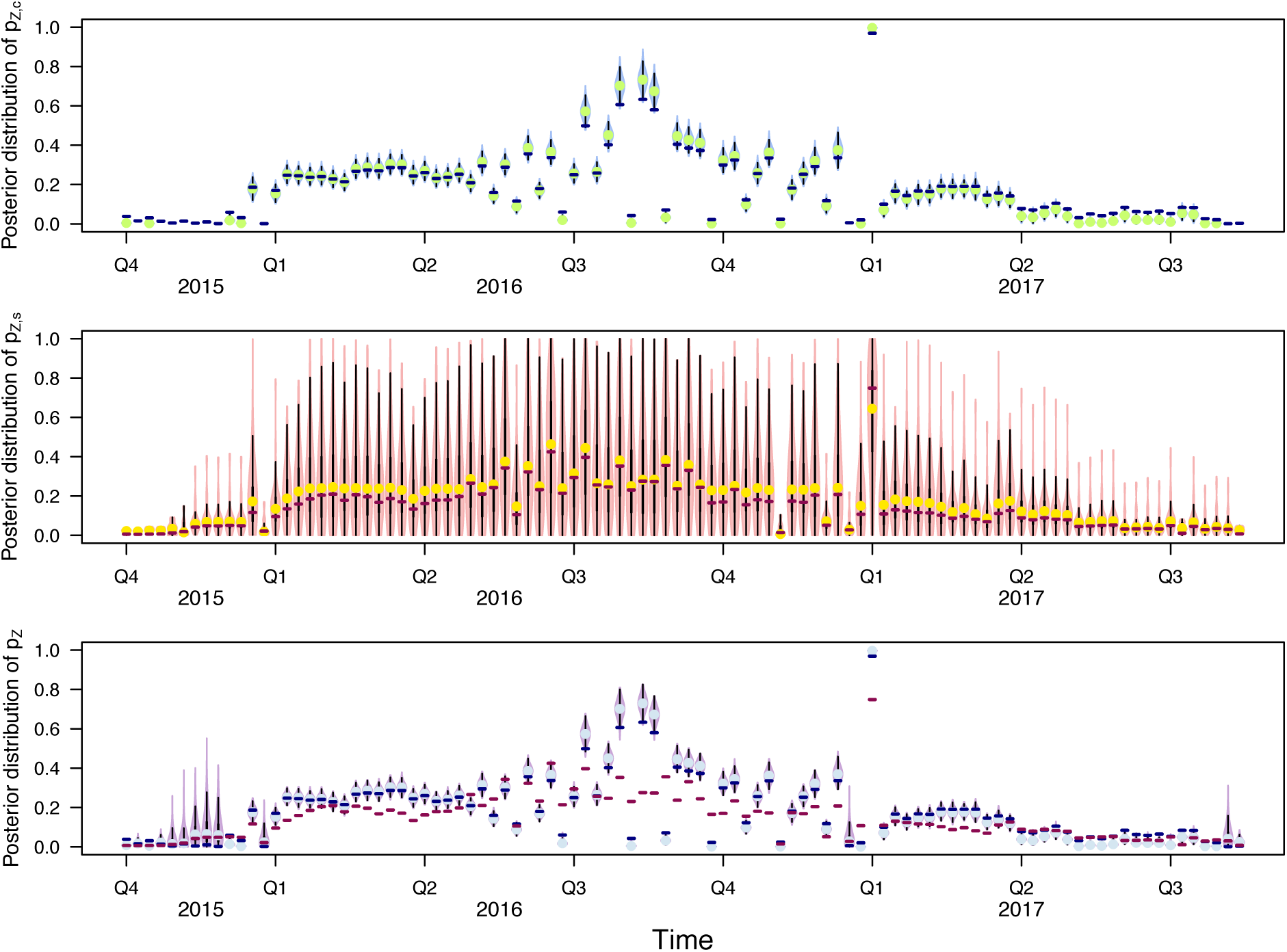
Posterior distributions of p_Z,c_ (top), p_Z,s_ (middle), and p_Z_ (bottom) for each time point using spatially aggregated data only. Top: Horizontal navy line indicates 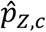. Middle: Horizontal maroon line indicates 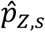. Bottom: Horizontal navy line indicates 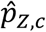 and horizontal maroon line indicates 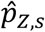.

**Figure S5.**
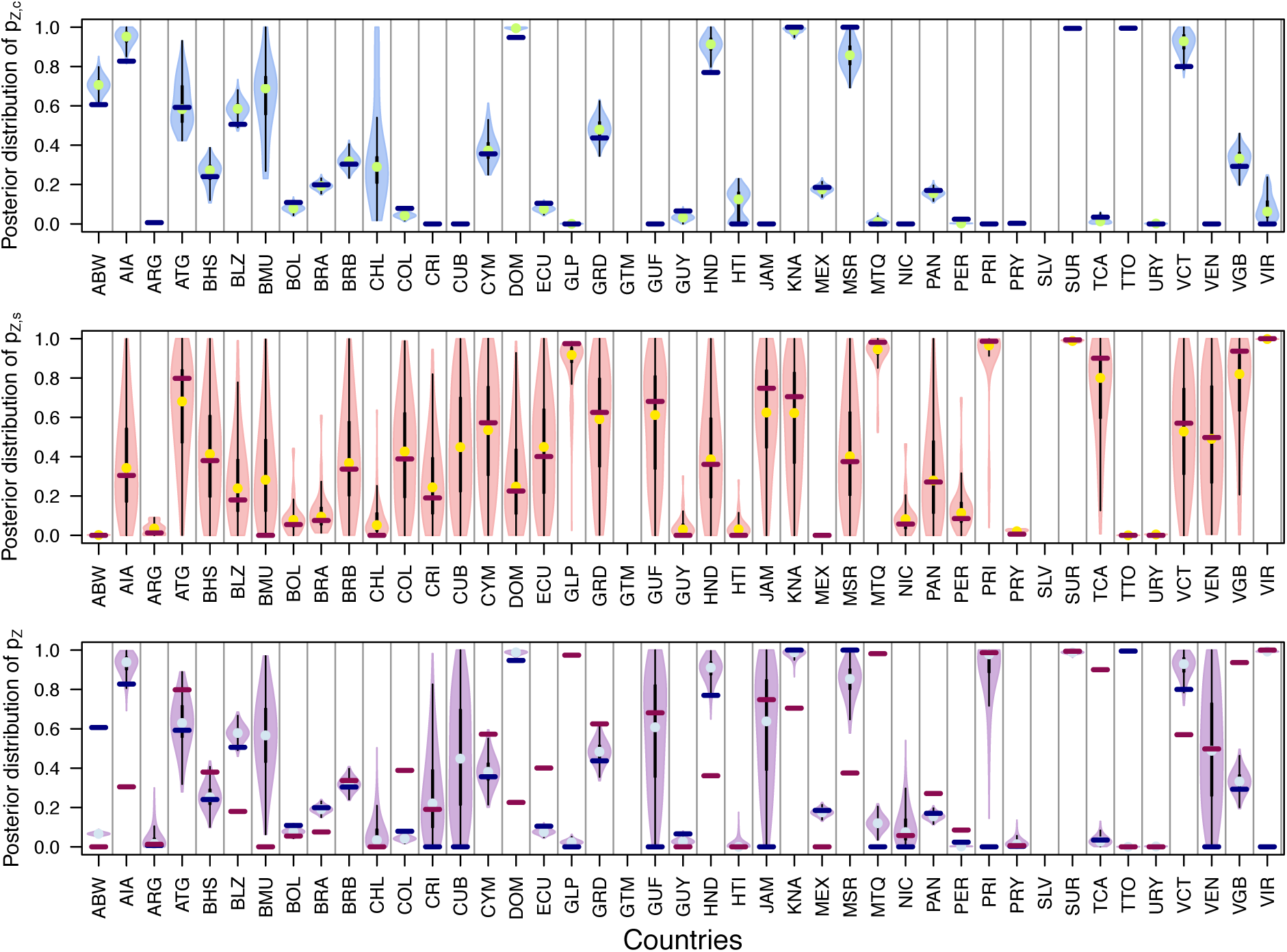
Posterior distributions of p_Z,c_ (top), p_Z,s_ (middle), and p_Z_ (bottom) for each country using cumulative data only. Top: Horizontal navy line indicates 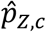. Middle: Horizontal maroon line indicates 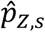. Bottom: Horizontal navy line indicates 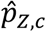 and horizontal maroon line indicates 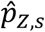.

## References

1. Hay SI, Snow RW. The Malaria Atlas Project: Developing Global Maps of Malaria Risk. PLOS Med. 2006 Dec 5;3(12):e473.

2. Murray CJL, Lopez AD. Mortality by cause for eight regions of the world: Global Burden of Disease Study. Lancet. 1997 May 3;349(9061):1269–76.

3. Bhatt S, Gething PW, Brady OJ, Messina JP, Farlow AW, Moyes CL, et al. The global distribution and burden of dengue. Nature. 2013/04/07. 2013 Apr 25;496(7446):504–7.

4. Tunbridge AJ, Breuer J, Jeffery KJM. Chickenpox in adults – Clinical management. J Infect. 2008;57(2):95–102.

5. Iroh Tam P-Y, Obaro SK, Storch G. Challenges in the Etiology and Diagnosis of Acute Febrile Illness in Children in Low- and Middle-Income Countries. J Pediatric Infect Dis Soc. 2016 Apr 7;5(2):190–205.

6. Caterino JM, Kline DM, Leininger R, Southerland LT, Carpenter CR, Baugh CW, et al. Nonspecific Symptoms Lack Diagnostic Accuracy for Infection in Older Patients in the Emergency Department. J Am Geriatr Soc. 2019 Mar 1;67(3):484– 92.

7. Mattar S, Tique V, Miranda J, Montes E, Garzon D. Undifferentiated tropical febrile illness in Cordoba, Colombia: Not everything is dengue. J Infect Public Health. 2017;10(5):507–12.

8. Hayes EB, Sejvar JJ, Zaki SR, Lanciotti RS, Bode A V., Campbell GL. Virology, pathology, and clinical manifestationsof West Nile Virus Disease. Emerg Infect Dis. 2005;11(8).

9. ten Bosch QA, Clapham HE, Lambrechts L, Duong V, Buchy P, Althouse BM, et al. Contributions from the silent majority dominate dengue virus transmission. PLOS Pathog. 2018 May 3;14(5):e1006965.

10. Majowicz SE, Hall G, Scallan E, Adak GK, Gauci C, Jones TF, et al. A common, symptom-based case definition for gastroenteritis. Epidemiol Infect. 2007/08/09. 2008 Jul;136(7):886–94.

11. Balogh EP, Miller BT, Ball JR. Improving diagnosis in health care. Washington, D.C.: The National Academies Press; 2015.

12. Banoo S, Bell D, Bossuyt P, Herring A, Mabey D, Poole F, et al. Evaluation of diagnostic tests for infectious diseases: general principles. Nat Rev Microbiol. 2006 Sep 1;4:S21.

13. Dong J, Olano JP, McBride JW, Walker DH. Emerging Pathogens: Challenges and Successes of Molecular Diagnostics. J Mol Diagnostics. 2008;10(3):185–97.

14. Pfaller MA. Molecular Approaches to Diagnosing and Managing Infectious Diseases: Practicality and Costs. Emerg Infect Dis J. 2001;7(2):312.

15. Heymann DL. T he international response to the outbreak of SARS in 2003. Philos Trans R Soc Lond B Biol Sci. 2004 Jul 29;359(1447):1127–9.

16. Rao PN, van Eijk AM, Choubey S, Ali SZ, Dash A, Barla P, et al. Dengue, chikungunya, and scrub typhus are important etiologies of non-malarial febrile illness in Rourkela, Odisha, India. BMC Infect Dis. 2019 Jul 3;19(1):572.

17. Opatowski L, Baguelin M, Eggo RM. Influenza interaction with cocirculating pathogens and its impact on surveillance, pathogenesis, and epidemic profile: A key role for mathematical modelling. PLoS Pathog. 2018 Feb 15;14(2):e1006770– e1006770.

18. WHO. WHO surveillance case definitions for ILI and SARI [Internet]. Available from: https://www.who.int/influenza/surveillance_monitoring/ili_sari_surveillance_case_definition/en/

19. Mayxay M, Castonguay-Vanier J, Chansamouth V, Dubot-Pérès A, Paris DH, Phetsouvanh R, et al. Causes of non-malarial fever in Laos: a prospective study. Lancet Glob Heal. 2013;1(1):e46–54.

20. Hochedez P, Jaureguiberry S, Debruyne M, Bossi P, Hausfater P, Brucker G, et al. Chikungunya infection in travelers. Emerg Infect Dis. 2006 Oct;12(10):1565–7.

21. Yactayo S, Staples JE, Millot V, Cibrelus L, Ramon-Pardo P. Epidemiology of Chikungunya in the Americas. J Infect Dis. 2016 Dec 5;214(suppl_5):S441–5.

22. Hills SL, Fischer M, Petersen LR. Epidemiology of Zika Virus Infection. J Infect Dis. 2017 Dec 16;216(suppl_10):S868–74.

23. Pacheco O, Beltrán M, Nelson CA, Valencia D, Tolosa N, Farr SL, et al. Zika Virus Disease in Colombia — Preliminary Report. N Engl J Med. 2016.

24. Waggoner JJ, Pinsky BA. Zika Virus: Diagnostics for an Emerging Pandemic Threat. Kraft CS, editor. J Clin Microbiol. 2016 Apr 1;54(4):860 LP–867.

25. Braga JU, Bressan C, Dalvi APR, Calvet GA, Daumas RP, Rodrigues N, et al. Accuracy of Zika virus disease case definition during simultaneous Dengue and Chikungunya epidemics. PLoS One. 2017;12(6):1–14.

26. PAHO. Zika virus infection: Data, maps, and statistics [Internet]. Available from: https://www.paho.org/hq/index.php?option=com_topics&view=rdmore&cid=8095&item=zika-virus-infection&type=statistics&Itemid=41484&lang=en

27. PAHO. Reported cases of dengue fever in the Americas [Internet]. Available from: https://www.paho.org/data/index.php/en/mnu-topics/indicadores-dengue-en/dengue-nacional-en/252-dengue-pais-ano-en.html

28. PAHO. Chikungunya [Internet]. Available from: https://www.paho.org/hq/index.php?option=com_topics&view=article&id=343&Itemid=40931&lang=en

29. WHO. Zika virus disease [Internet]. Available from: https://www.who.int/csr/disease/zika/case-definition/en/

30. van der Walt S, Schonberger J, Nunez-Iglesias J, Boulogne F, Warner J, Yager N, et al. scikit-image: Image processing in Python. PeerJ. 2014;2:e453.

31. Oliphant T. A guide to NumPy. In: 1st ed. Trelgol Publishing USA; 2006.

32. Charrel R, Mögling R, Pas S, Papa A, Baronti C, Koopmans M, et al. Variable Sensitivity in Molecular Detection of Zika Virus in European Expert Laboratories: External Quality Assessment, November 2016. McAdam AJ, editor. J Clin Microbiol. 2017;55(11):3219–26.

33. Hobbs NT, Hooten MB. Bayesian Models: A Statistical Primer for Ecologists. Princeton University Press; 2015. 320 p.

34. R Development Core Team. R: A Language and Environment for Statistical Computing. R Found Stat Comput Vienna Austria. 2016;0:{ISBN} 3-900051-07-0.

35. Lourenço J, Maia de Lima M, Faria NR, Walker A, Kraemer MUG, Villabona-Arenas CJ, et al. Epidemiological and ecological determinants of Zika virus transmission in an urban setting. Jit m, editor. Elife. 2017;6:e29820.

36. Perkins TA, Metcalf CJE, Grenfell BT, Tatem AJ. Estimating Drivers of Autochthonous Transmission of Chikungunya Virus in its Invasion of the Americas. PLoS Curr. 2015;7.

37. Mina M, Guterman L, Omer S. Zika in the United States: Dynamics, Transmission, and Strategies for Control. Am J Clin Pathol. 2018 Jan 11;149(suppl_1):S202– S202.

38. Rodriguez-Barraquer I, Costa F, Nascimento EJM, Nery N, Castanha PMS, Sacramento GA, et al. Impact of preexisting dengue immunity on Zika virus emergence in a dengue endemic region. Science (80-). 2019;363(6427):607–10.

39. Vogels CBF, Rückert C, Cavany SM, Perkins TA, Ebel GD, Grubaugh ND. Arbovirus coinfection and co-transmission: A neglected public health concern?PLOS Biol. 2019 Jan 22;17(1):e3000130.

40. Bell D, Wongsrichanalai C, Barnwell JW. Ensuring quality and access for malaria diagnosis: how can it be achieved?Nat Rev Microbiol. 2006;4(9):682–95.

41. Apat DO, Gachohi JM, Karama M, Kiplimo JR, Sachs SE. Temporal variation in confirmed diagnosis of fever-related malarial cases among children under-5 years by community health workers and in health facilities between years 2013 and 2015 in Siaya County, Kenya. Malar J. 2017;16(1):454.

42. Soljak M, Samarasundera E, Indulkar T, Walford H, Majeed A. Variations in cardiovascular disease under-diagnosis in England: national cross-sectional spatial analysis. BMC Cardiovasc Disord. 2011;11(1):12.

43. Freitas LP, Cruz OG, Lowe R, Sá Carvalho M. Space–time dynamics of a triple epidemic: dengue, chikungunya and Zika clusters in the city of Rio de Janeiro. Proc R Soc B Biol Sci. 2019 Oct 9;286(1912):20191867.

44. Borchering RK, Huang A, Mier-y-Teran-Romero L, Rojas DP, Rodriguez-Barraquer I, Katzelnick LC, et al. Dengue after Zika: characterizing impacts of Zika emergence on endemic dengue transmission. bioRxiv. 2019;

45. Ximenes RA de A, Miranda-Filho D de B, Brickley EB, Montarroyos UR, Martelli CMT, Araújo TVB de, et al. Zika virus infection in pregnancy: Establishing a case definition for clinical research on pregnant women with rash in an active transmission setting. PLoS Negl Trop Dis. 2019 Oct 7;13(10):e0007763.

46. Brady OJ, Osgood-Zimmerman A, Kassebaum NJ, Ray SE, de Araújo Vem, da Nóbrega AA, et al. The association between Zika virus infection and microcephaly in Brazil 2015–2017: An observational analysis of over 4 million births. PLOS Med. 2019 Mar 5;16(3):e1002755.

47. Carlson CJ, Mendenhall E. Preparing for emerging infections means expecting new syndemics. Lancet. 2019;394(10195):297

